# Neuroanatomical Considerations for Optimizing Thalamic Deep Brain Stimulation in Tourette Syndrome

**DOI:** 10.1101/2020.09.29.20200501

**Authors:** Takashi Morishita, Yuki Sakai, Hitoshi Iida, Saki Yoshimura, Atsushi Ishii, Shinsuke Fujioka, Saori C. Tanaka, Tooru Inoue

## Abstract

**Background:** Deep brain stimulation (DBS) of the centromedian (CM) thalamic nucleus has reportedly been used to treat severe Tourette syndrome (TS) with promising outcomes; however, it remains unclear how DBS electrode position and stimulation parameters modulate the specific area and related networks. We aimed to evaluate the relationships between the anatomical location of stimulation fields and clinical responses including therapeutic and side effects.

**Methods:** We collected data from eight TS patients treated with DBS. We evaluated the clinical outcomes using Yale Global Tic Severity Scale (YGTSS), Yale-Brown Obsessive Compulsive Scale (Y-BOCS), and Hamilton Depression Rating Scale (HAM-D). The DBS lead location was evaluated in the normalised brain space using a 3-D atlas. The volume of tissue activated (VTA) was determined, and the associated normative connective analyses were performed to link the stimulation field with the therapeutic and side effects.

**Results:** The mean follow-up period was 10.9 ± 3.9 months. All clinical scale showed significant significant improvement. While the VTA associated with therapeutic effects covers the CM and ventrolateral nuclei and showed association with motor networks, those associated with paraesthesia and dizziness were associated with stimulation of the ventralis caudalis and red nucleus, respectively. Depressed mood was associated with the spread of stimulation current to the mediodorsal nucleus and showed association with limbic networks.

**Conclusion:** Our study addresses the importance of accurate implantation of DBS electrodes for obtaining standardised clinical outcomes and suggests that meticulous programming with careful monitoring of clinical symptoms may improve outcomes.

## Introduction

Tourette syndrome (TS) is a neuropsychiatric disorder characterised by involuntary tic movements and psychiatric comorbidities. Although symptoms are subtle and subside spontaneously in childhood, some patients affected by this syndrome suffer from severe, debilitating involuntary movements. Common treatment options for TS include medical and behavioural therapies, and deep brain stimulation (DBS) may be indicated for the treatment of severe TS. Since the publication of the first report on the successful application of DBS therapy for TS [1], reports on its clinical outcomes have been published [2, 3]. A previous study concerning the treatment of TS using DBS has reported mixed results although overall clinical outcomes were favourable [2-4]. Additionally, a recent long-term follow-up study showed that a loss of benefit occurs after several years of continuous thalamic stimulation [5].

Several targets for DBS therapy including the centromedian (CM) thalamic nucleus, globus pallidus interna (GPi), and anterior limb of internal capsule have been proposed for the treatment of TS using DBS, and stimulation of these targets is reportedly equally effective [2, 3]. However, response to the therapy varies among patients. The heterogeneity of the therapeutic effects is potentially associated with the employed patient selection criteria, surgical technique, and DBS programme. A potential factor associated with DBS failure is lead misplacement [6], and a recent study reported that the incidence of DBS lead misplacement in Parkinson’s disease cohort was higher than 15% [7]. Even though the incidence of lead misplacement in the TS cohort has not been reported, we suspect its occurrence in some patients with TS refractory to DBS in the reported cohorts.

Because the visualisation of each thalamic nucleus for stereotactic targeting has been challenging and because neurophysiological microelectrode recordings (MER) have not been established for the CM nucleus, it is difficult to determine the occurrence of lead misplacement in a patient with TS showing poor DBS outcome. Additionally, detailed subdivisions of the thalamic structures associated with favourable stimulation outcome have not been identified as in the case of the GPi or subthalamic nucleus, which are stimulated for the treatment of Parkinson’s disease although a multi-centre analysis reported preferred stimulation points [8]. Clinicians should be aware that a subtle difference in the lead position and electrically stimulated areas results in the activation of different neuronal networks; however, it remains unclear how DBS electrode position and stimulation parameters modulate the specific area and related networks.

The position of DBS electrodes is conventionally reported using Cartesian coordinates relative to the midcommissural point, but recent studies have addressed the importance of considering anatomical variations while determining the optimal DBS electrode position [9]. To analyse patient-specific electrode positions, various authors have used a common population-based standard Montreal Neurological Institute (MNI) space [8, 10], and recent studies have reported the utility of three-dimensional anatomical atlases and structural connectome [11, 12]. Using these neuroimaging techniques, we aimed to evaluate the relationships between the anatomical location of implanted DBS leads and clinical responses. We also evaluated stimulation-induced side effects as they interfere with optimising stimulation settings for symptoms suppression.

## Methods

### Study Design

We prospectively recorded the clinical course of the patients and reviewed the charts to obtain detailed information. To interpret the DBS lead positions associated with clinical benefits and stimulation-induced adverse events, we evaluated the DBS lead position in the postoperative images. This study was approved by our institutional review board (IRB) named Fukuoka University-Medical Ethics Review Board (IRB approval number: U02-02-001), and informed consent was obtained from the participants. This study was conducted in accordance with the Declaration of Helsinki.

We included eight patients who underwent DBS surgery at our department, and all patients completed at least a six-month follow-up. DBS therapy was indicated for patients with severe, medication-refractory symptoms who were older than 12 years of age. One patient (case 7) underwent DBS surgery because of severe cervical tic with the risk of spinal cord injury despite relatively low clinical scoring owing to the absence of phonic tics and other comorbidities. All patients underwent multidisciplinary evaluation performed by a team consisting of a neurologist, psychiatrist, paediatrician, and neurosurgeon, as recommended by a recent guidelines paper [13]. The demographics of patients are summarised in Table 1.

**Table 1.**
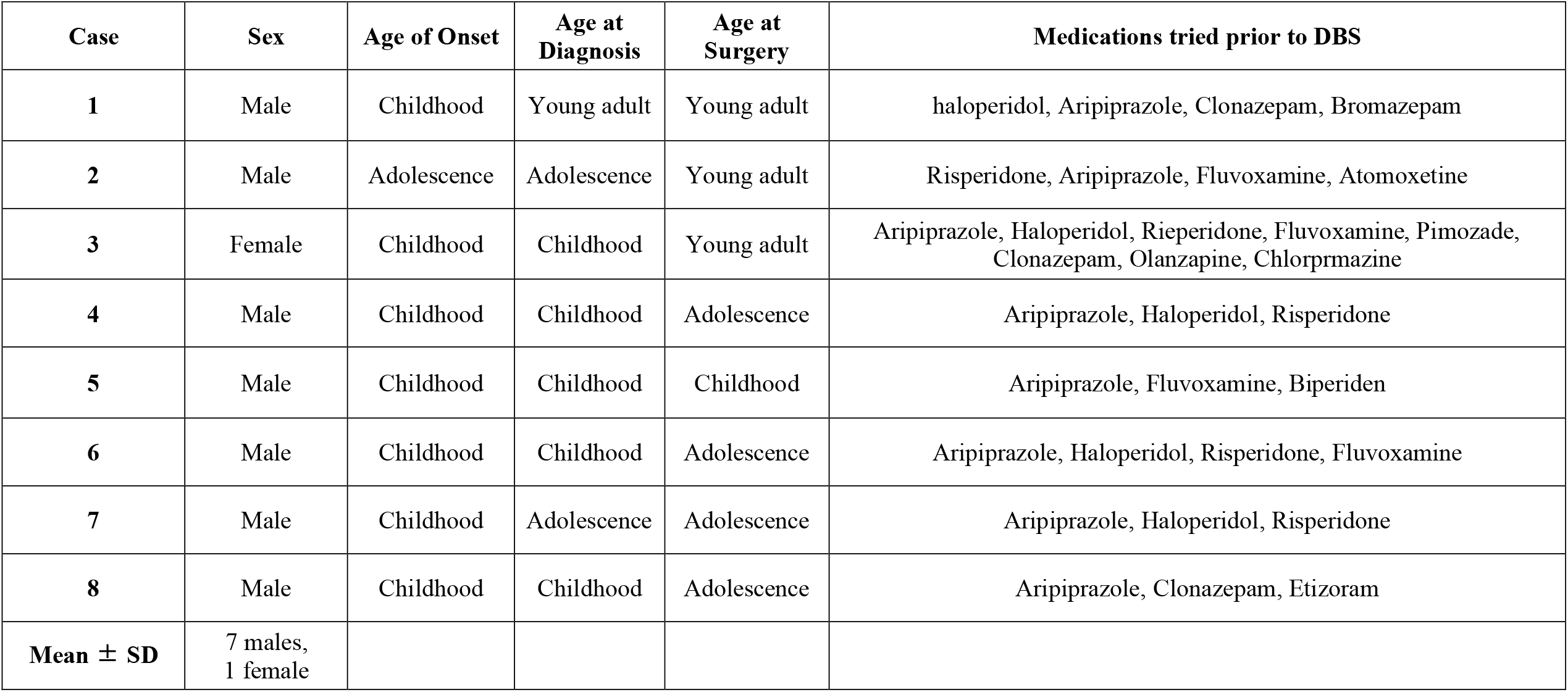
Patient Demographics. **Childhood: age 3-12; Adolescence: age 13-19, Young adult: age 20-39** **DBS = deep brain stimulation; SD = standard deviation**

We evaluated the clinical outcomes using Yale Global Tic Severity Scale (YGTSS) [14], Yale-Brown Obsessive Compulsive Scale (Y-BOCS) [15], and Hamilton Depression Rating Scale (HAM-D) [16] (Table 2). These evaluations were performed prior to surgery and six months and one year after surgery. Additionally, we recorded the adverse events associated with surgery and neurostimulation. Mood changes were recorded in the medical records on the basis of self-reported complaints of the patients.

**Table 2.** Clinical outcomes. Abbreviations. F/U = follow up; HAM-D = Hamilton Depression Rating Scale; N/A = not applicable; Y-BOCS = Yale-Brown Obsessive Compulsive Scale; YGTSS = Yale Global Tic Severity Scale.

|  |  | YGTSS Severity |  |  | YGTSS Impairment |  |  | Y-BOCS |  |  | HAM-D |  |  |
| --- | --- | --- | --- | --- | --- | --- | --- | --- | --- | --- | --- | --- | --- |
| Case | F/U period | Baseline | 6 mo | Last F/U | Baseline | 6 mo | Last F/U | Baseline | 6 mo | Last F/U | Baseline | 6 mo | Last F/U |
| 1 | 12 | 40 | 20 | 16 | 50 | 30 | 30 | 23 | 21 | 19 | 10 | 8 | 6 |
| 2 | 12 | 50 | 23 | 12 | 50 | 30 | 30 | 32 | 4 | 0 | 17 | 9 | 12 |
| 3 | 12 | 32 | 15 | 14 | 40 | 20 | 10 | 0 | 0 | 0 | 19 | 14 | 10 |
| 4 | 18 | 47 | 19 | 15 | 50 | 20 | 20 | 30 | 0 | 7 | 4 | 1 | 5 |
| 5 | 12 | 50 | 43 | 27 | 50 | 40 | 20 | 15 | 15 | 15 | 1 | 2 | 2 |
| 6 | 9 | 48 | - | 23 | 50 | - | 20 | 19 | - | 8 | 11 | N/A | 2 |
| 7 | 6 | 25 | 9 | 9 | 50 | 10 | 10 | 10 | 0 | 0 | 3 | 1 | 1 |
| 8 | 6 | 43 | 12 | 10 | 40 | 10 | 10 | 5 | 0 | 0 | 4 | 0 | 0 |
| Mean ± SD | 10.9 ± 3.9 | 41.9 ± 9.1 | 20.1 ± 11.2 | 15.8 ± 6.3 | 47.5 ± 4.6 | 22.9 ± 11.1 | 18.8 ± 8.3 | 16.8 ± 11.5 | 5.7 ± 8.7 | 6.1 ± 7.5 | 8.6 ± 6.7 | 5.0 ± 5.4 | 4.8 ± 4.4 |
| P-values | N/A | N/A | 0.018 | 0.012 | N/A | 0.017 | 0.010 | N/A | 0.043 | 0.028 | N/A | 0.028 | 0.035 |

### Surgical Procedure

Because we have reported our DBS procedures previously [17, 18], we describe our method briefly herein. Magnetic resonance imaging (MRI) was performed on all the patients for stereotactic planning using an MRI scanner (Ingenia 1.5T, Philips, Netherland), and the MRI sequences included volumetric T1-weighted imaging (T1WI) with contrast, volumetric fast grey matter acquisition T1 inversion recovery (FGATIR), and diffusion tensor imaging. Using commercialised software (iPlan stereotaxy, Brainlab, Germany), we identified the target structure and planned the trajectory of DBS lead implantation. Above mentioned MRI sequences were automatically fused, and the anterior commissure (AC), posterior commissure (PC), and the midline plane to anchor the Cartesian coordinate system were determined. The tentative target was initially set at 5 mm lateral and 4 mm posterior to the midcommissural point on the AC-PC plane. The trajectory was planned such that the DBS lead would not injure the blood vessels, lateral ventricle, and sulci during T1WI with contrast, and the target (presumed lead tip position at the ventral surface of the CM nucleus) was modified according to the location of the CM nucleus in each case. The CM nucleus was identified as a relatively high-intensity area on FGATIR images, as reported previously [18]. The target was meticulously checked using the mammillothalamic tract and the red nucleus (RN) as landmarks (Supplementary Figure 1).

In this study, all patients underwent simultaneous bilateral implantation of DBS leads (model 3387, Medtronic, Minnesota) and implantation of an implantable pulse generator (IPG) (Activa RC, Medtronic, Minnesota) on the same day. The first four patients underwent DBS lead implantation under local anaesthesia; however, three of them could not maintain the fixed head position during the awake procedure. Thus, we used general anaesthesia throughout the surgical procedure for all system implantations, including the stereotactic frame fixation for the remaining four cases (cases 5–8).

### DBS Programming

Electrical stimulation was delivered immediately following DBS system implantation in the first six patients and two weeks after DBS lead implantation in the remaining two patients (cases 7 and 8). The initial stimulation was delivered using the monopolar setting, which activates the second most ventral contacts (contacts 1 and 9) bilaterally, with the following parameters: amplitude, 2.0 volts or milliamperes; pulse width (PW), 60 microseconds; and frequency, 130 Hz.

One month after surgery, we measured the threshold levels of stimulation-induced side effects in each contact at a fixed PW of 60 microseconds and frequency of 130 Hz. We carefully interviewed the patients at each programming session, and we increased the stimulation intensity within the therapeutic window such that acute side effects could be avoided. At each programming visit, new programming parameters were saved as a new group of settings so that patients can go back to the previous setting in case they experience the chronic stimulation-induced side effects such as mood changes. If the clinical response was insufficient with maximum intensity at one contact, we activated another contact to apply interleaving stimulations using multiple contacts. The frequency of programming depends on the patients’ accessibility and the clinical response to the stimulation.

### DBS Lead Localization

Stereotactic computed tomography (CT) scans were performed on all patients to determine the lead location, typically on postoperative day nine at the point when the pneumocephalus was resolved. The CT image was fused to a preoperative scan to measure the DBS lead trajectory and contact positions relative to the midcommissural point in each case. We compared the stereotactic planning and DBS lead locations to measure the stereotactic errors. Stereotactic targeting error was calculated as the distance between the planned target point and trajectory of the implanted lead [19].

All imaging data were preprocessed using the Lead-DBS software (www.lead-dbs.org) [20]. Briefly, all (pre- and postoperative) CT and MRIs were linearly co-registered to the preoperative T1WI using SPM (https://www.fil.ion.ucl.ac.uk/spm/software/spm12/). Registration between postoperative CT and preoperative T1WI was further refined using the linear registration within the subcortical target region of interest to minimise non-linear bias caused by the surgery [21]. All data were normalised into the standard MNI space. This procedure was performed with the whole-brain non-linear SyN registration implemented in Advanced Normalisation Tools (ANTs, http://stnava.github.io/ANTs/) [22] using the “effective (low variance)” setting with subcortical refinement as implemented in Lead-DBS [20]. Electrode trajectories and contacts were automatically pre-reconstructed using PaCER algorithm [23] and manually refined using Lead-DBS. We also determined the lead trajectory and contact positions using subject-independent 3D atlases of the thalamic nuclei [24]. We considered that contacts existed within those thalamic nuclei when spherical regions of 0.2 mm radius around contacts overlapped with each nucleus: CM, mediodorsal (MD), ventral lateral (VL), and ventral posterior (VP) nuclei. Detected electrodes and thalamic nuclei were visualised using Lead-DBS.

### Volume of Tissue Activated Mapping

We calculated the volume of tissue activated (VTA) under the conditions with the optimised stimulation parameters (Table 3) and those with stimulation-induced side effects (Table 3, 4). All VTA calculations were conducted using Lead-DBS [25]. A volume conductor model was constructed on the basis of a tetrahedral volume mesh that included the DBS electrode and surrounding tissues. Conductivities of 0.14 S/m were assigned to grey and white matter [26]. On the basis of the volume conductor model, the electric field distribution was simulated using the FieldTrip-SimBio pipeline that was integrated into Lead-DBS (https://www.mrt.uni-jena.de/simbio/index.php/; http://fieldtriptoolbox.org). The electric field distribution was thresholded for magnitudes above a commonly used value of 0.2 V/mm [27, 28] to define the extent of VTA.

**Table 3.** The stimulation parameters at last follow up and those induced mood changes. All side effects were reversal by reducing the stimulation intensity. *Interleaving stimulation settings were applied. Abbreviation. PW = pulse width

|  |  | Left |  |  |  |  | Right |  |  |  |  |  |
| --- | --- | --- | --- | --- | --- | --- | --- | --- | --- | --- | --- | --- |
| Case | Time Point | Cathode | Anode | PW (μsec) | Frequency (Hz) | Amplitude (V or mA) | Cathode | Anode | PW (μsec) | Frequency (Hz) | Amplitude (V or mA) | Chronic Stimulation-induced Side Effects |
| 1 | 12 mo* | 0 | Case | 60 | 100 | 3.8 V | 9 | Case | 60 | 100 | 3.8 V | None |
|  |  | 2 | Case | 60 | 100 | 2.5 V | 11 | Case | 60 | 100 | 2.5 V |  |
|  | 2 mo | 0 | Case | 120 | 125 | 3.0 V | 9 | Case | 120 | 125 | 3.5 V | Depressed mood. |
| 2 | 12 mo | 2 | Case | 90 | 130 | 4.0 mA | 10 | Case | 90 | 130 | 4.0 mA | None |
|  | 11 mo | 1 | Case | 90 | 125 | 4.0 V | 9 | Case | 90 | 125 | 3.9 V | Depressed mood.<br>Aggressiveness.<br>Slurred speech. |
| 3 | 12 mo* | 1 | Case | 90 | 110 | 2.5 V | 9 | Case | 90 | 110 | 2.0 V | None |
|  |  | 3 | Case | 90 | 110 | 3.0 V | 11 | Case | 90 | 110 | 3.0 V |  |
|  | 11 mo | 2 | Case | 90 | 130 | 3.5 mA | 11 | Case | 90 | 130 | 2.9 mA | Depressed mood and anxiety. |
| 4 | 18 mo | 3 | 1 | 100 | 125 | 3.5 mA | 11 | 9 | 100 | 125 | 3.5 mA | None |
|  | 8 mo* | 0 | Case | 90 | 125 | 4.2 V | 9 | Case | 90 | 125 | 4.2 V | Depressed mood and suicidal ideation. |
|  |  | 2 | Case | 120 | 125 | 4.0 V | 11 | Case | 120 | 125 | 4.0 V |  |
|  | 14 mo | 2 | Case | 100 | 130 | 4.0 mA | 10 | Case | 100 | 130 | 4.0 mA | Depressed mood. |
| 5 | 12 mo* | 1 | Case | 60 | 125 | 2.8 mA | 9 | Case | 60 | 125 | 2.8 mA | None |
|  |  | 2 | Case | 120 | 125 | 3.0 mA | 10 | Case | 120 | 125 | 3.0 mA |  |
| 6 | 9 mo* | 2 | Case | 100 | 125 | 3.2 mA | 10 | Case | 100 | 125 | 3.2 mA | None |
|  |  | 3 | Case | 100 | 125 | 4.0 mA | 11 | Case | 100 | 125 | 4.0 mA |  |
| 7 | 6 mo* | 2 | Case | 90 | 125 | 3.1 mA | 10 | Case | 90 | 125 | 3.1 mA | None |
|  |  | 3 | Case | 90 | 125 | 3.0 mA | 11 | Case | 90 | 125 | 3.0 mA |  |
| 8 | 6 mo | 2 | Case | 90 | 130 | 3.8 mA | 10 | Case | 90 | 130 | 4.2 mA | None |

**Table 4.** Threshold levels of acute stimulation-induced side effects. Threshold levels of stimulation-induced side effects in each contact were measure at a fixed PW of 60 microseconds and frequency of 130 Hz. N/A = not applicable

| Case | Side | Contact 0 |  | Contact 1 |  | Contact 2 |  | Contact 3 |  |
| --- | --- | --- | --- | --- | --- | --- | --- | --- | --- |
|  |  | Threshold Level | Side Effect | Threshold Level | Side Effect | Threshold Level | Side Effect | Threshold Level | Side Effect |
| 1 | Left | 5.8 V | Dizziness | 7.2 V | Dizziness | N/A | None | N/A | None |
|  | Right | 3.5 V | Dizziness | 6.0 V | Dizziness | 8.9 V | Paresthesia in the left hand | N/A | None |
| 2 | Left | 1.8 mA | Dizziness | 4.0 mA | Tinnitus on the left ear | 6.6 mA | Paresthesia in the right hand | N/A | None |
|  | Right | 4.0 mA | Dizziness | 5.0 mA | Paresthesia in the left forearm | 8.2 mA | Paresthesia in the left fingers | N/A | None |
| 3 | Left | 1.8 mA | Dizziness | 3.6 mA | Dizziness | 6.5 mA | Dizziness | N/A | None |
|  | Right | 2.5 mA | Dizziness | 2.9 mA | Dizziness | 3.1 mA | Paresthesia in the left hand | 4.4 mA | Dizziness |
| 4 | Left | 5.5 V | Nausea | N/A | None | 7.0 V | Nausea | 6.7 V | Nausea |
|  | Right | 3.2 V | Electric shock sensation in the head | 4.9 V | Paresthesia in the left hand and face | 6.2 V | Anxiety | 7.8 V | Paresthesia in the back of the neck |
|  | Left (post-revision) | 3.0 mA | Nausea | 3.6 mA | Paresthesia in the right hand, Squeezing sensation of the head | 4.8 mA | Deep tactile sensation in the right ear | 8.4 mA | Squeezing sensation of the head |
| 5 | Left | 5.1 mA | Dizziness, H/A | 7.4 mA | Dizziness, H/A | N/A | None | N/A | None |
|  | Right | 6.8 mA | Dizziness, H/A | N/A | None | N/A | None | N/A | None |
| 6 | Left | 6.0 mA | Dizziness | 6.5mA | Dizziness | N/A | None | N/A | None |
|  | Right | 6.5 mA | Dizziness | N/A | None | N/A | None | N/A | None |
| 7 | Left | 2.6 mA | Dizziness | 3.6 mA | Dizziness, Paresthesia in the right hand | 6.2 mA | “Funny sensation” in the head | 7.5 mA | Paresthesia in the right hand |
|  | Right | 4.5 mA | Dizziness | 5.9 mA | Paresthesia in both forearms | 7.0 mA | Dizziness | N/A | None |
| 8 | Left | 5.8 mA | Dizziness | N/A | None | N/A | None | N/A | None |
|  | Right | 6.0 mA | Dizziness | N/A | None | N/A | None | N/A | None |

### Normative Connectome

To characterise the brain connectome differences from very adjacent VTAs and to elucidate the brain-wide mechanism of therapeutic stimulation and side effects, we used the population-averaged atlas of the macroscale human structural connectome derived from diffusion-weighted imaging (DWI) data (N = 842, Human Connectome Project) [29]. Considering the limited volume of data, the VTAs in the right hemisphere were non-linearly transformed into those in the left hemisphere using ANTs, and all VTAs were pooled across hemispheres. We created specific VTAs that did not overlap with the other VTAs and depicted the 300 fibres from them using DSI studio (http://dsi-studio.labsolver.org). All detected structural fibres were visualised using Lead DBS. We evaluated the proportion of the 300 fibres that passed through each brain region defined by Harvard-Oxford cortical/subcortical atlases [30] combined with the cerebellum from AAL atlas [31]. All small parcels of the cerebellum defined in AAL were integrated into one binarized parcel. The proportion of 300 fibres was thresholded above 0.02 and visualised using a circular plot.

### Statistical Analysis

To compare the pre- and post-DBS clinical scores on YGTSS, Y-BOCS, and HAM-D, we used Wilcoxon signed rank test. Statistical analyses were performed using SPSS version 21.0 (IBM Corp., Armonk, NY, USA), and p < 0.05 indicated statistical significance.

## Results

### Clinical Outcomes

The mean follow-up period was 10.9 ± 3.9 months, but one patient (case 6) missed a formal 6-month follow-up owing to the occurrence of the COVID-19 pandemic during the study period. YGTSS severity and impairment scores improved from 41.9 ± 3.9 and 47.5 ± 4.6 at the baseline to 15.8 ± 6.3 (z = −2.52, p = 0.012) and 18.8 ± 8.3 (z = −2.59, p = 0.010) at the last follow-up, respectively. Furthermore, Y-BOCS and HAM-D scores improved from 16.8 ± 11.5 and 8.6 ± 6.7 at the baseline to 6.1 ± 7.5 (z = −2.20, p = 0.028) and 4.8 ± 4.4 (z = −2.11, p = 0.035) at the last follow-up, respectively. Clinical outcomes are summarised in Table 2. It is noteworthy that one patient (Case 8) experienced an enormous microlesion effect while the tic movements had disappeared for a week after surgery.

The surgical adverse events included wound dehiscence and lead misplacement. Two patients (cases 1 and 4) experienced wound dehiscence of the scalp incision site and underwent a wound revision. One patient (case 4) underwent a left DBS lead revision owing to insufficient phonic tic suppression despite the motor tic improvement.

### DBS Lead Location

Coordinates of the preoperative stereotactic targeting and the measured DBS lead locations are summarised in Supplementary Table 1. Stereotactic targeting error was 1.2 ± 0.5 mm. Analysis of DBS lead positions indicated that 15 of 17 DBS leads, including a misplaced lead, penetrated the CM nucleus and that 11 leads penetrated the MD nucleus (Supplementary Table 2). The dorsal contacts of the quadripolar electrodes were located in the ventrolateral nucleus in all cases except for the initial left lead of case 4. In case 4, the tip of the misplaced DBS lead on the left was on the border between MD and CM nuclei, and three contacts were positioned in the MD nucleus. Two ventral contacts and two dorsal contacts of the revised lead in case 4 were successfully placed in the CM nucleus and ventrolateral nucleus, respectively (Supplementary Table 2 and Supplementary Movie 1). The DBS lead contact locations in the normalised brain space are shown in Fig. 1 and Supplementary Movie 1.

**Figure 1.**
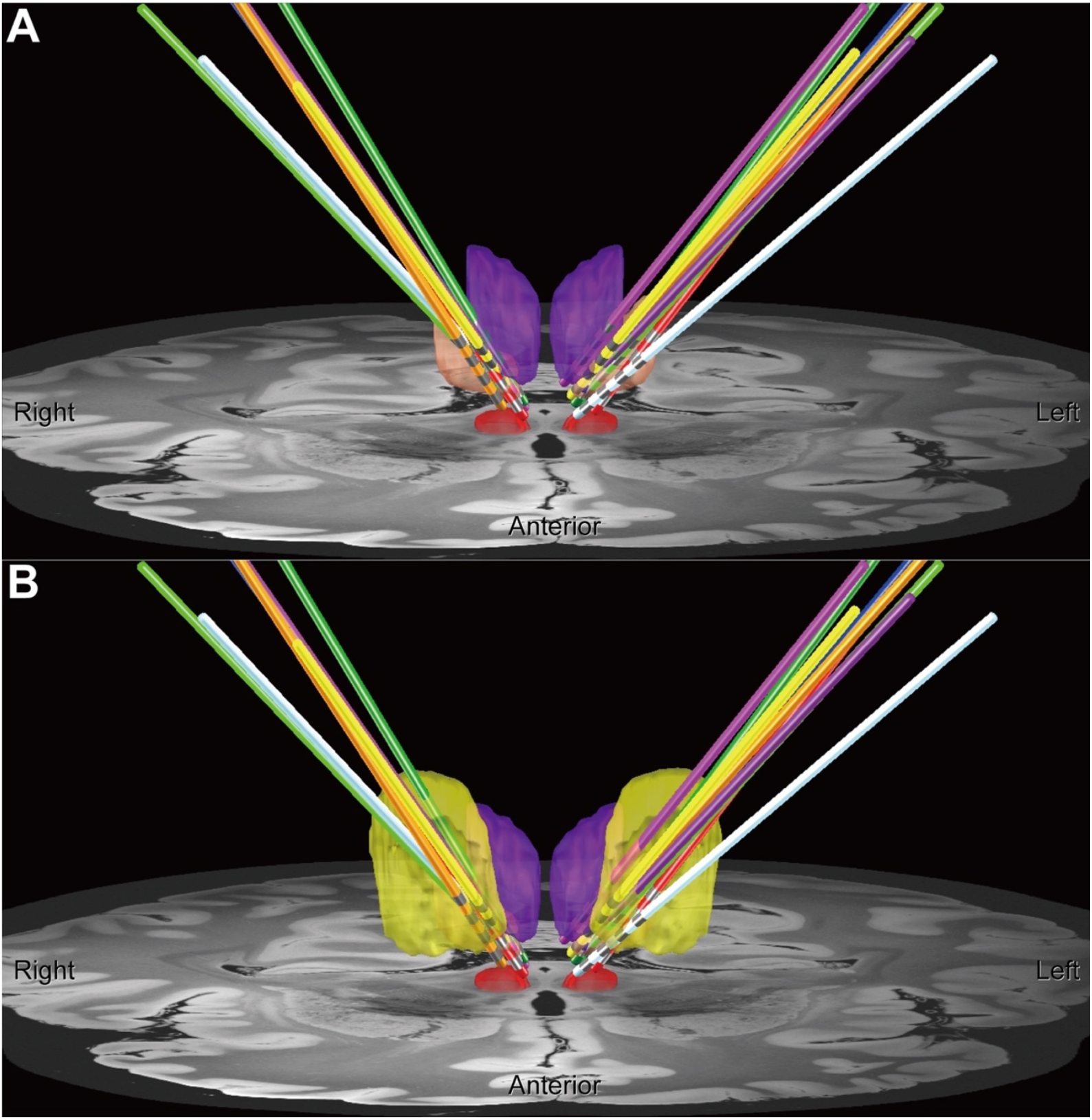
Lead electrodes placement in MNI space. Lead electrodes with the CM nucleus (peach), RN (red), MD nucleus (purple), and ventral lateral dorsal/ventral nucleus (yellow). In (A), the ventral lateral dorsal/ventral nucleus was removed. The lead electrodes of each patient were displayed in different colours (case 1, orange; 2, red; 3, blue; 4, purple; 4 after the repositioning, dark purple (only in the left hemisphere); 5, light green; 6, yellow; 7, light blue: 8, green).

### Clinical Response According to the Stimulated Area

Contacts 2 (10) and 3 (11) were likely to be selected as the active contacts (Table 3). Therapeutic effects were achieved by stimulation of the border between the CM and VL nuclei in our case series. Depressed mood or anxiety were reported after chronic stimulation (Table 3). Four patients commonly experienced mood changes after chronic stimulation for several days after a programming session. The programming parameters at the last follow-up and chronic side effects are summarised in Table 3. The common acute side effects were dizziness and paraesthesia in the contralateral upper extremity of the active DBS lead. Dizziness and paraesthesia tended to be observed with high-intensity stimulation of the two ventral contacts and two middle contacts, respectively. These acute stimulation-induced side effects are summarised in Table 4.

### Stimulated Areas and Normative Connectome

In Fig. 2, we show the VTAs related to the therapeutic stimulation and side effects. We found the VTA related to the clinical response in the border between CM and VL nuclei (blue area in Fig. 2A). Although the paraesthesia-related region mostly overlapped the therapeutic region, it extended slightly into lateral, medial, and anterior directions (green area). The VTA related to dizziness extended into the ventral direction and overlapped with the RN (orange area). We found that chronic stimulation of the relatively medial and dorsal region entering the MD nucleus led to a depressed mood (purple area).

**Figure 2.**
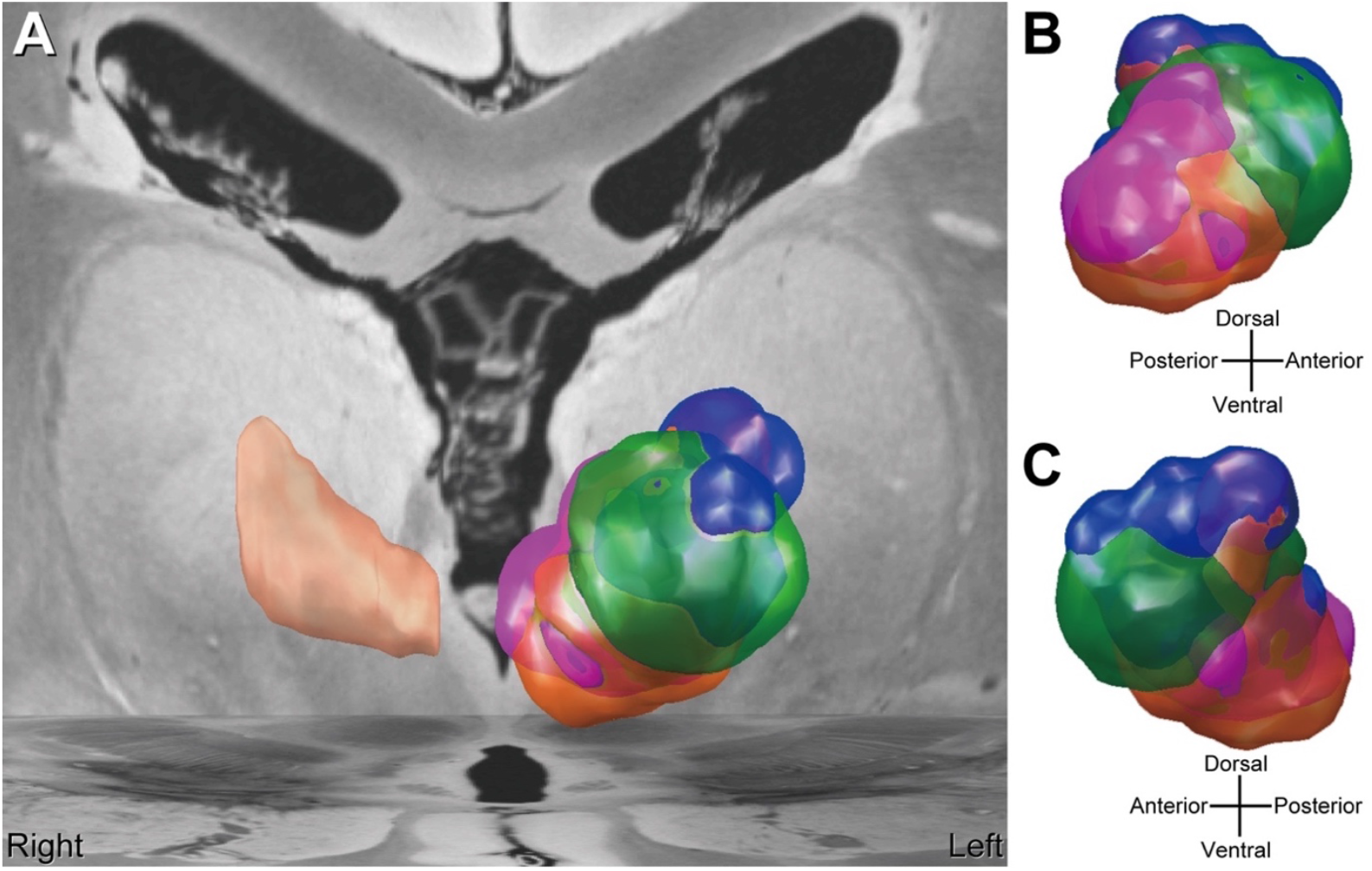
VTAs related to therapeutic stimulation and side effects. Each colour of VTAs represents areas associated with the following effects: blue = therapeutic effect, orange = dizziness, green = paraesthesia, and purple = depressed mood. The peach-coloured region in the right hemisphere is the CM nucleus. (A) front, (B) medial, and (C) lateral view.

Using the normative connectome, we found that each VTA related to the therapeutic stimulation and side effects showed clearly different network properties (Fig. 3). Brain regions connected with each VTA are summarised using the circular plot (Fig. 4, Supplementary Figure 2). Fibres of therapeutic stimulation were characterised by more dense connections with the precentral gyrus than were those of side effects (Fig. 4A). Dizziness- and paraesthesia-related VTAs extended fibres into relatively specific brain regions (Fig. 4B, 4C). In contrast, the stimulation-induced mood change was related to more spatially distributed brain regions (Fig. 4D). Specifically, we detected dizziness related fibres into the cerebellorubral network (Fig. 3B, 4B). The paraesthesia symptom was characterised by the fibres that connect the thalamus and insular cortex (Fig. 3C, 4C). We found a relatively dense connection with the thalamus in the amygdala and the orbitofrontal cortex, which are related to depressed mood (Fig. 3D, 4D).

**Figure 3.**
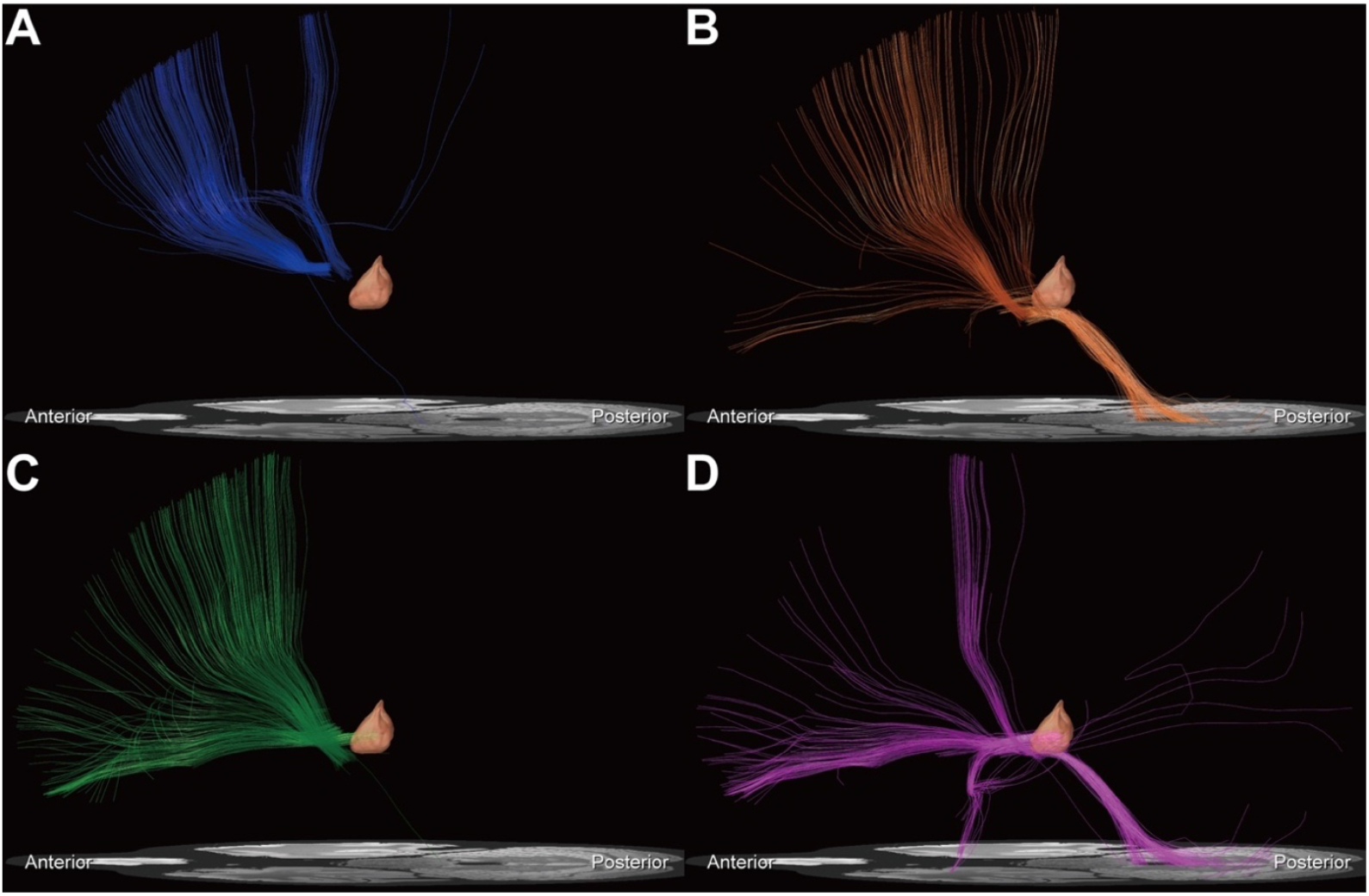
Normative connectome from VTAs related to therapeutic stimulation and side effects. The normative connectome from VTAs related to the (A) therapeutic stimulation, (B) dizziness, (C) paraesthesia, and (D) depressed mood. The peach-coloured region is the CM nucleus.

**Figure 4.**
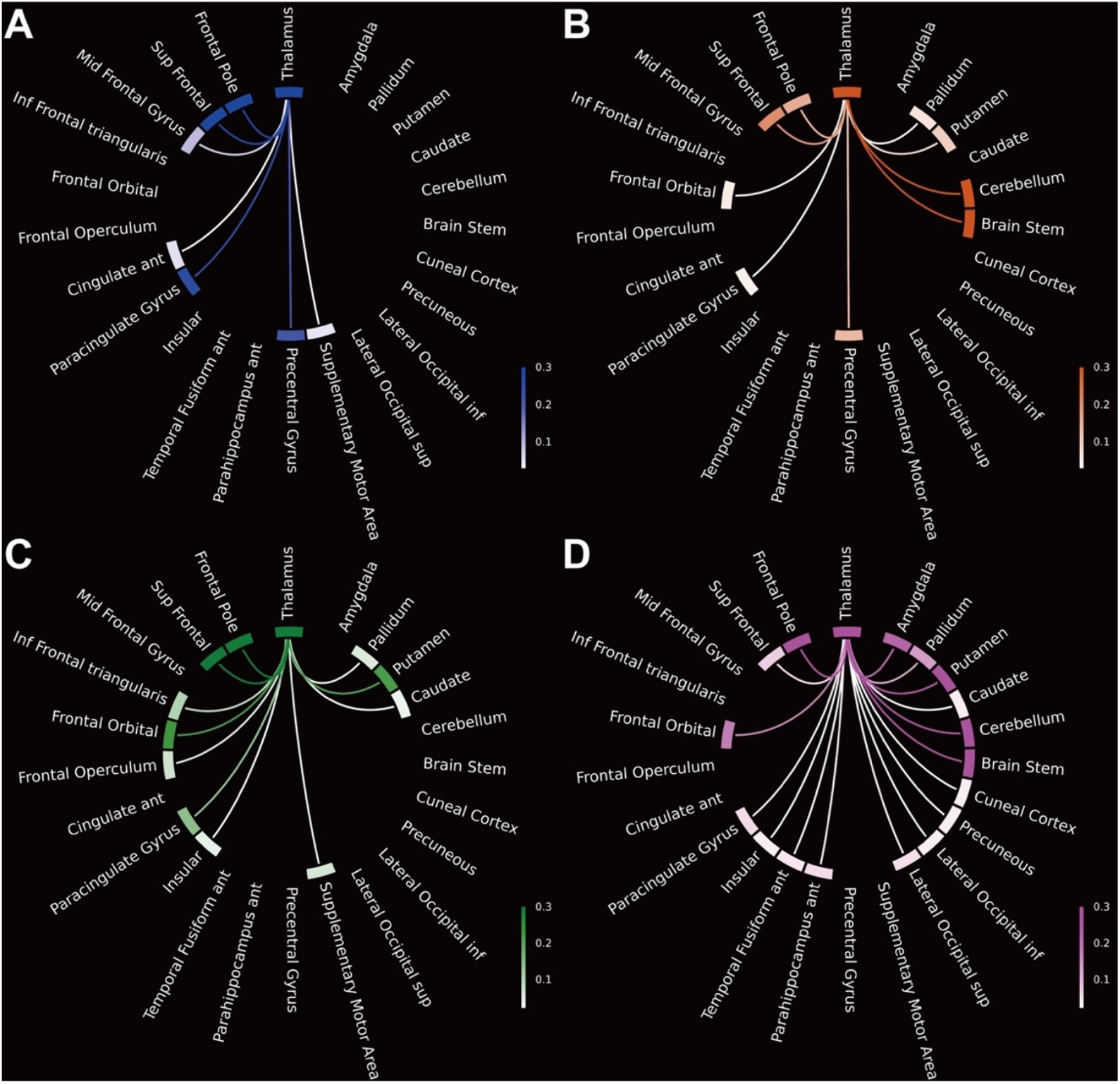
Proportion of the structural fibres projected from VTAs related to therapeutic stimulation and side effects. The circular plot represents the proportion of projected fibres that connect the thalamus and the other brain regions which have at least one connectivity (see Supplementary Figure 2 for all brain regions); (A) therapeutic stimulation, (B) dizziness, (C) paraesthesia, and (D) depressed mood. Abbreviations: ant = anterior; inf = inferior; mid = middle; post = posterior; sup = superior.

## Discussion

Studies that evaluated clinical outcomes of DBS in patients with TS have reported mixed results on a case-by-case basis although overall outcomes were favourable [2-4], and our results were consistent with those of the past reports. The results of our study showed a variety of DBS contacts (Fig. 1, Supplementary Movie 1) and illustrated the relationship between the DBS lead locations and clinical effects inclusive of therapeutic and side effects (Fig. 2, 3, 4).

Regarding the stereotactic planning technique, the lead trajectory is determined according to the safety issues [32]. Indirect targeting using the same template coordinates is not appropriate as this technique does not consider the anatomical variations in the subcortical structures [9]. According to the literature, DBS leads were presumed to pass through the ventral-oralis complex (Vo) nucleus to the CM nucleus; however, our series showed that there were a variety of lead locations even among the responders (Fig. 1, Supplementary Movie 1). We consider that such differences may have led to the occurrence of difference in threshold levels of stimulation-induced side effects (Table 3 and 4). Various factors associated with the surgical procedure such as brain shift and distortion of the frame are considered to deviate the lead position from the planned target [19]. Although we did not measure the electrode positions intraoperatively, an image-guided procedure using intraoperative CT or MRI may improve the stereotactic accuracy and precision [33, 34].

A potential factor predicting the incidence of a favourable outcome is the microlesion effect, and this issue also underpins the importance of the accurate lead implantation. Microlesion effect has been reported to be associated with the optimal lead position in other movement disorders such as Parkinson’s disease and essential tremor [35, 36]. Similarly, we have noted a temporary but complete resolution of tic movements immediately after surgery in a patient (case 8). These findings indicate that stimulation of a specific area is associated with tic suppression. In this context, clinical outcomes of TS DBS may be improved with accurate and precise DBS lead implantation in the specific area. Additionally, our experience that DBS lead repositioning improved the clinical outcome (case 4) addresses the importance of implanting DBS leads in the optimal position to maximise the benefit.

Difficulty in DBS programming for TS exists, in that, immediate response may not be observed in the clinical setting unlike that in the case of Parkinson’s disease or essential tremor. Practitioners programming the DBS, however, should understand the relationship between the DBS lead and the surrounding structures. A subtle difference in stimulating position can lead to various effects through different properties of the brain network as the thalamus is quite a dense structure of many small nuclei that relays information between different brain regions [24]. The VTA and normative connectivity analyses linked our clinical findings with how electrical currents spreads to the surrounding neuroanatomical structures according to the situations (Fig. 2, 3, 4).

The VTA associated with therapeutic effects covered the dorsal area of the CM nucleus and the VL nucleus, and our normative connectome analysis showed its association with the motor networks (Fig. 3A, 4A). This finding is consistent with that reported by a recent study showing that the relationship of tic reduction with the fibres connecting the thalamus and the motor cortex [37]. On the contrary, it is well known that high electrical intensity is usually required for tic suppression [4], and therapeutic stimulation may also modulate the limbic systems sub-clinically inducing psychiatric side effects.

With regard to the immediate side effects, the ventralis caudalis (Vc) nucleus and RN are located posterolateral and ventral to the CM nucleus, respectively. Spread of stimulation current to the Vc nucleus and RN may induce paraesthesia and dizziness, respectively. These findings were partly supported by the findings of normative connectome analyses. These side effects can be observed immediately when the stimulation intensity is above the threshold levels in the clinical setting, but mood changes are likely to be detected several days after increasing the stimulation intensity. The lower threshold levels of these stimulation-induced side effects may indicate lead misplacement, and clinicians should be aware that the careful evaluation of clinical response to each stimulation parameter is important to estimate the lead location.

The reported mood changes are considered to result from the use of supra-threshold intensity for the electrical stimulation of the limbic network, and this finding was supported by that of the normative connectome analysis, that is, the electrically activated area associated with the mood change is connected to limbic structures such as the amygdala (Fig. 3D, 4D). We consider that the high intensity of electrical current to the MD nucleus was associated with the side effects, and this may underpin the mechanism of loss of benefits due to side effects such as reduced levels of energy following long-term thalamic DBS reported recently [5]. In our study, the depressed moods were temporary because we decreased the stimulation intensity or changed active contacts when patients experienced the side effects. However, chronic DBS may induce irreversible effects as reported recently [38], and chronic high-intensity stimulation of the MD nucleus may result in irreversible mood changes or loss of beneficial effects.

Although our study has demonstrated promising clinical outcomes of DBS therapy for severe, medication-refractory TS and provides a guide for identifying stimulation areas in the thalamus that yield desirable effects, it has several limitations. The evaluation was not blinded, and records of mood change in our study were based on patients’ reports rather than on quantified data. To confirm our findings, multi-centre studies with a higher number of patients are warranted. A registry collecting meticulous data from multiples centres may address these issues as reported by several authors [3, 8, 39].

## Conclusion

Our study addresses the importance of accurate implantation of DBS electrodes for obtaining standardised clinical outcomes and suggests that meticulous programming with careful monitoring of clinical symptoms may improve outcomes. Clinicians should attempt to detect any subtle changes in the clinical symptoms at each clinical visit for better stimulation adjustment, and in this context, findings of our study may be useful with regard to the systematic programming paradigm. Further meticulous evaluation of clinical effects of neurostimulation at specific area with a large study population is warranted.

## Data Availability

Data Availability
The clinical outcomes of our cohort were registered to the International Tourette Deep Brain Stimulation Database and Registry (https://tourettedeepbrainstimulationregistry.ese.ufhealth.org/). The personal data including the imaging data obtained from the study subjects will not be distributed openly to protect the patients' privacy. We used the population-averaged atlas of the macroscale human structural connectome derived from DWI (http://dsi-studio.labsolver.org/). For imaging analysis, we used free software inclusive of ANTs (http://stnava.github.io/ANTs/) (Avants et al., 2008), Lead-DBS (https://www.lead-dbs.org/) (Horn et al., 2019), PaCER algorithm (Husch et al., 2018), FieldTrip-SimBio (https://www.mrt.uni-jena.de/simbio/index.php/; http://fieldtriptoolbox.org), and DSI studio (http://dsi-studio.labsolver.org). The subject-independent 3D atlases of the thalamic nuclei are also openly available online (http://www.humanmotorthalamus.com/) (Ilinsky et al., 2018).

## Abbreviations

AC: Anterior commissure
ANTs: Advanced Normalisation Tools
CM: centromedian
DBS: Deep brain stimulation
DWI: Diffusion-weighted imaging
FGATIR: fast grey matter acquisition T1 inversion recovery
GPi: globus pallidus interna
IPG: Implantable pulse generator
MNI: Montreal Neurological Institute
MRI: Magnetic resonance imaging
PC: Posterior commissure
PW: Pulse width
RN: Red nucleus
TS: Tourette syndrome
VTA: Volume of tissue activated
YGTSS: Yale Global Tic Severity Scale

## Acknowledgements

None

